# Investigating Uptake and Impact of Genetic and Genomic Evaluation Following Perinatal Demise

**DOI:** 10.64898/2026.04.22.26347546

**Authors:** Kara Mossler, Etta Genevieve D’Orazio, Katherine Hall, Kathryn Osann, Virginia Kimonis, Fabiola Quintero-Rivera

**Author notes:** **Corresponding authors:** - Kara Mossler, Division of Genetics and Genomic Medicine, Division of Neonatology, Department of Pediatrics, University of California, Irvine, Irvine, CA, USA, - Virginia Kimonis, Division of Genetics and Genomic Medicine, Department of Pediatrics, University of California, Irvine, Irvine, CA, USA. **Funding Statement:** This research received no specific grant from any funding agency in the public, commercial, or not-for-profit sectors. **Data Availability Statement:** The data that support the findings of this study are available from the corresponding author upon reasonable request.

## Abstract

**Objective:** The decline of the perinatal demise rate is slowing and demises are often unexplained. Significant research has been done regarding diagnostic yield and genetic causes of demise, but little is known about how Geneticist involvement impacts outcomes. The goal of the study was to evaluate post-mortem genetic testing practices and effects of the geneticist’s involvement.

**Methods:** Retrospective data from 111 perinatal demise cases was examined, including rates of prenatal genetic counseling, post-delivery genetics consult, genetic testing, and autopsy investigation.

**Results:** In this cohort 54% received genetic testing and 25% received a genetics consultation. When compared to those without, cases with genetic specialist involvement were associated with significant increases in testing uptake (p=0.007), diagnostic yield (p<0.001), and patient education (p<0.001). Second trimester stillbirths and those with fewer ultrasound (US) abnormalities were less likely to receive genetic testing (both p values <0.001) and consults (p<0.001, p=0.020).

**Conclusion:** Although ascertainment bias cannot be ruled out, this data demonstrates that geneticist involvement correlates with a higher rate of testing, greater diagnostic yield, and more thorough counseling. These findings underscore the importance of integrating genetics providers into perinatal postmortem healthcare teams.

**What is already known about this topic?:**

- Causes of perinatal demise often are undiagnosed, but genetic and congenital anomalies are common.
- ACOG recommends genetic testing for all perinatal demises

**What does this study add?:**

- Genetic testing is under-offered and should be offered more frequently.
- Genetic specialist involvement is associated with increased patient education, genetic testing uptake, and diagnostic yield
- Time and access to genetic specialists may drive testing rate
- Non-English language may be associated with decreased consultation rate

## Introduction

The American Academy of Pediatrics’ Definition II of perinatal demise includes fetal deaths/stillbirths (spontaneous abortions or intrauterine fetal demise, SAB-IUFD) at a gestational age of 20 weeks or more and infant deaths that occur on or before 28 days of life.^1^ This includes deaths before and during delivery, but does not include elective terminations. Thanks to improvements in newborn screening, public health programs, and access to prenatal care, the United States saw a steady decline in perinatal deaths through the 20th and 21^st^ centuries. In 1930, the SAB-IUFD rate was 37.8 per 1000 live births.^2^ This declined significantly each decade until 1990 and began to plateau thereafter (Figure 1). The decrease in neonatal deaths has been maintained at a marginally greater pace than that of SAB-IUFDs, dropping from 7.7 in 1982 to 3.0 in 2022 (61%).^2,3,4^ Unfortunately, the etiology of perinatal deaths often (in 24% of cases) goes unexplained.^5^ Recent national data indicated that birth defects were the third most common known cause of stillbirth and the second most common known cause of neonatal death.^5^ With so many incompletely explained cases, uncovering underlying genetic causes has huge potential to affect the way perinatal medicine is practiced. Genetic diagnoses also enable calculation of recurrence risk for future pregnancies, inform discussions regarding preconception and prenatal genetic testing, reveal treatment options, guide cascade testing for family members, and provide meaningful emotional closure to families.^6, 7^

**Figure 1.**
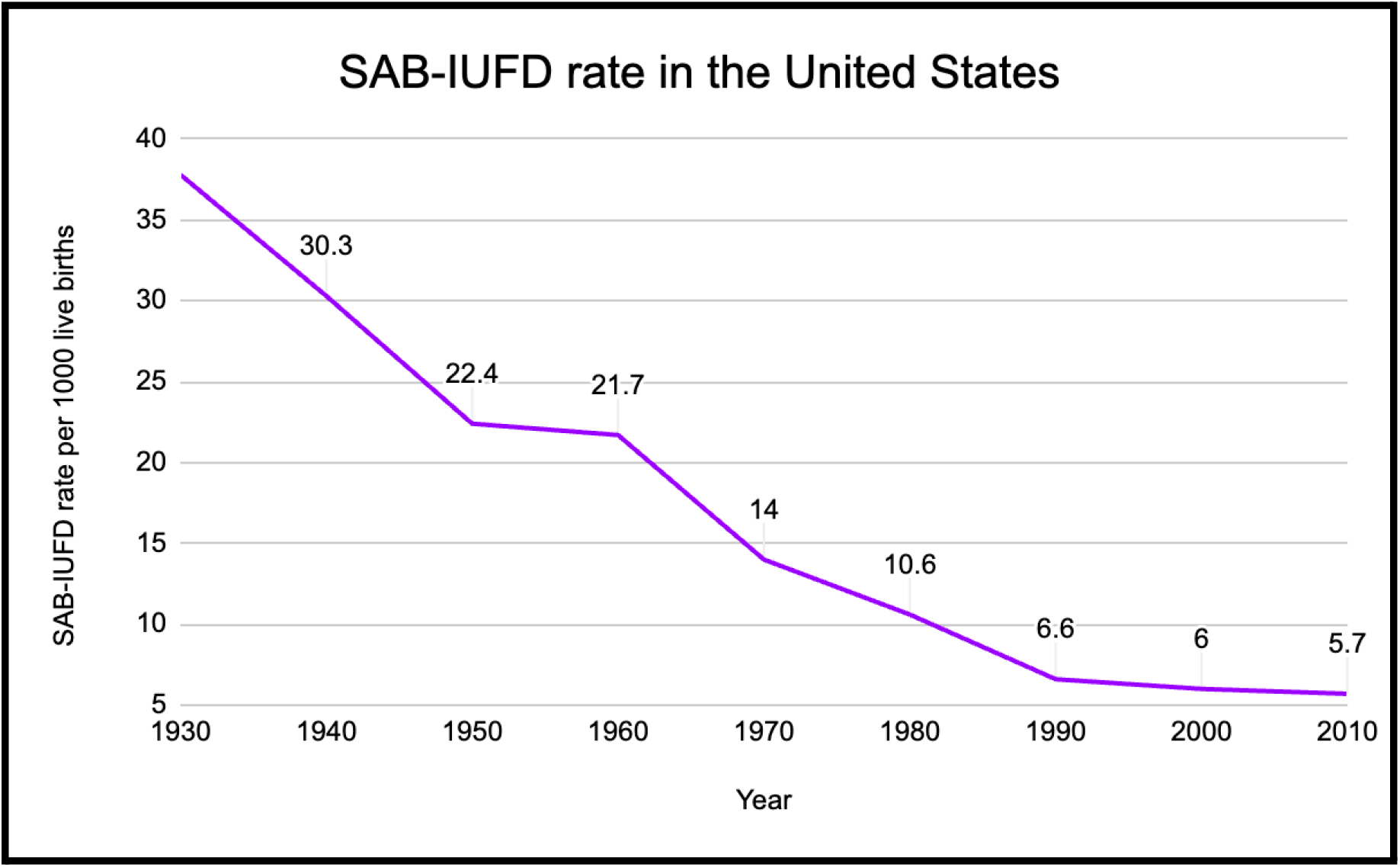
Rate of spontaneous abortions or intrauterine fetal demise (SAB-IUFDs) in the United States.^2,3,4^

The American College of Obstetricians and Gynecologists (ACOG) currently recommends autopsy, maternal evaluation, placental pathology, and genetic analysis as standard of care in all perinatal demise cases.^8^ Unfortunately, not all cases are offered genetic testing.^8, 9^ Determining which tests should be first line (e.g. chromosome analysis (karyotype), exome/genome studies (ES/GS)) is challenging and depends on the individual’s phenotype, but with rates of autopsy on the decline globally, genetic testing has and will become even more important to offer as a less invasive diagnostic option. In fact, some have argued for replacement of conventional autopsy with genetic autopsy altogether as the gold standard for diagnosis in perinatal demise.^10,11^ This study’s aim is to examine factors influencing the rates and yield of genetic testing and genetics consultation as they become more important than ever in management of perinatal demise.

## Methods

A chart review of perinatal demises that occurred at the University of California, Irvine Medical Center (UCIMC) from November 1, 2017, to December 1, 2021 was conducted. Perinatal demise records were paired with adult gestational parent records aged 18 years and older. Therapeutic (in the absence of SAB-IUFD) and elective abortions were excluded. Patients who had IVF (in vitro fertilization) were included in the study.

Records were identified through the electronic medical record (EMR) via the Department of Quality and Patient Safety. Delivery room and neonatal death records were also obtained through the EMR via the California Maternal Quality Care Collaborative. For data analysis purposes, delivery room deaths (within 12 hours of birth, no NICU care) were included in the total count of neonatal deaths. Data reviewed included gestational parent and fetal demographics, prenatal genetic counseling information, first anatomy US, autopsy documentation, post-delivery genetics consult documentation, and the result of genetic testing. The comparison analyses included all genetic testing received prenatally and postnatally by deceased perinates. Gestational parent income was estimated utilizing data based on ZIP code from the United States Census Bureau’s 2016-2020 American Community Survey.^12^ Number of points (out of six) discussed during genetic counseling sessions were reviewed in the documentation. These six counseling points were adapted from “Guidelines for Writing Letters to Patients” and include: description of significant histories (e.g. developmental, family) and pertinent test results, review of physical exam and/or diagnosis, natural history of condition, explanation of inheritance, summary of risk assessment, and cascade/parental testing recommended.^13^

Descriptive and inferential analyses were performed using the IBM Statistical Package for the Social Sciences (SPSS) Statistics version 28 (IBM SPSS Statistics for Macintosh, Armonk, NY, USA, IBM Corp). Descriptive analyses were used for demographics of gestational parents and deceased perinates. Univariate analysis was performed using Pearson’s chi-square tests or Fisher’s exact tests. A p-value of less than or equal to 0.05 was considered statistically significant. Due to the small number of individual racial and ethnic groups in the study, race and ethnicity were categorized as: White Non-Hispanic, Hispanic, and Other (included individuals of Asian, Black or African American, and multiracial descent). Similarly, due to the small size of preferred language groups, preferred languages were categorized into English or Other languages, which included Spanish, Vietnamese, Arabic, and Portuguese. Because a larger number of parents declared Christian religions, religious affiliation for gestational parents was categorized into Catholic, other Christian, other non-Christian, and non-religious.

## Results

### 3.1 Demographics and demise type

A total of 66 cases of neonatal death and 45 cases of SAB-IUFD met inclusion criteria (111 total). The most common races/ethnicities were Hispanic white (41.4%) and non-Hispanic white (23.4%). The language preferred by families was English for 81.1% followed by Spanish at 15.3%. Medicaid was the most common insurance (64%). Mean gestational age was 27 weeks, median was 26 weeks, and range was 20-41 weeks at birth. There was no statistically significant association between demise type and gestational age at delivery (p=0.222, Figure 2). See Table 1 in the Supplemental Material for detailed maternal demographics.

**Figure 2.**
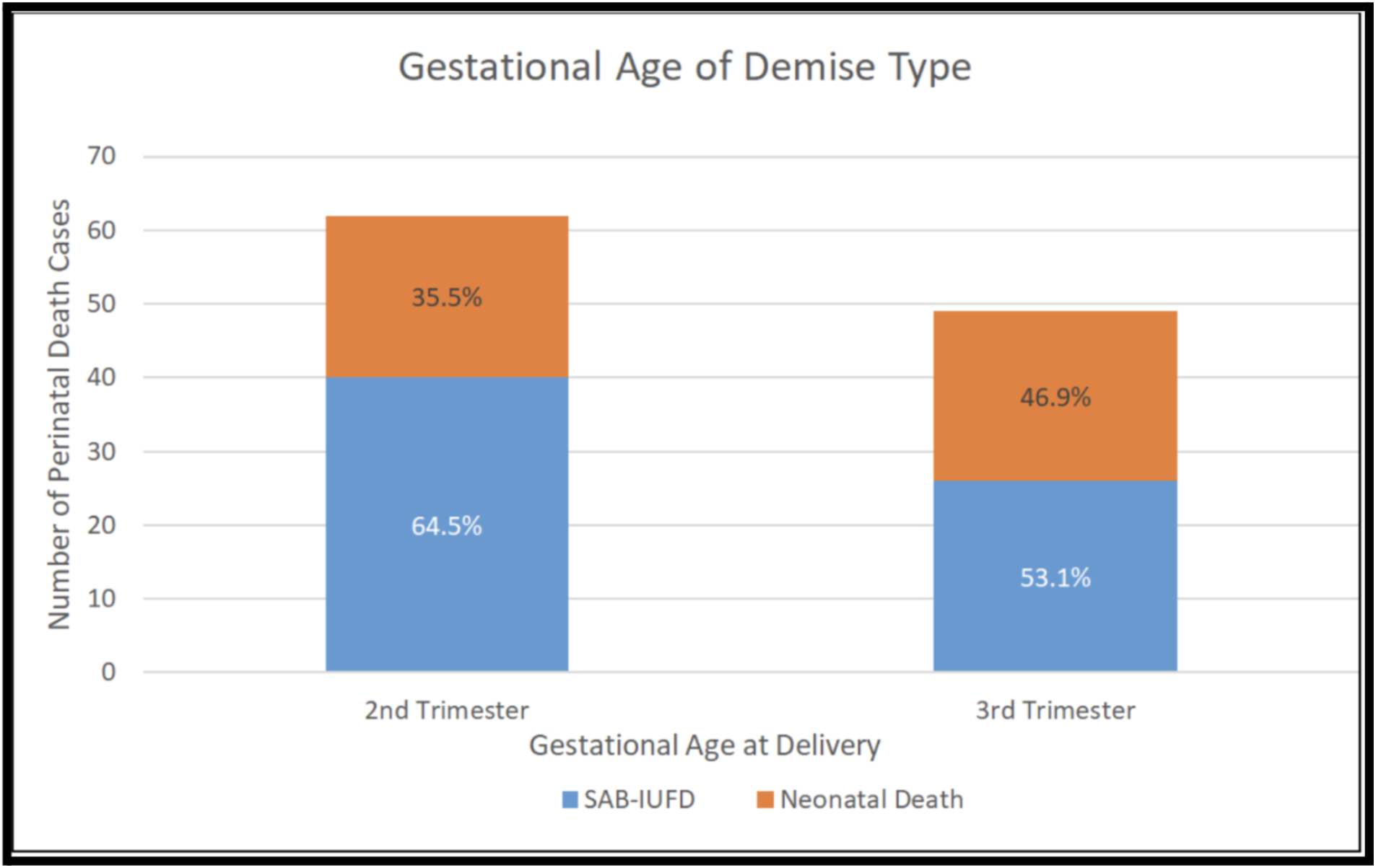
Comparison of gestational age at delivery with demise type (N=11). Second trimester (20 weeks 0 days to 27 weeks 6 days). Third trimester (28 weeks 0 days onward).

**Table 1:**
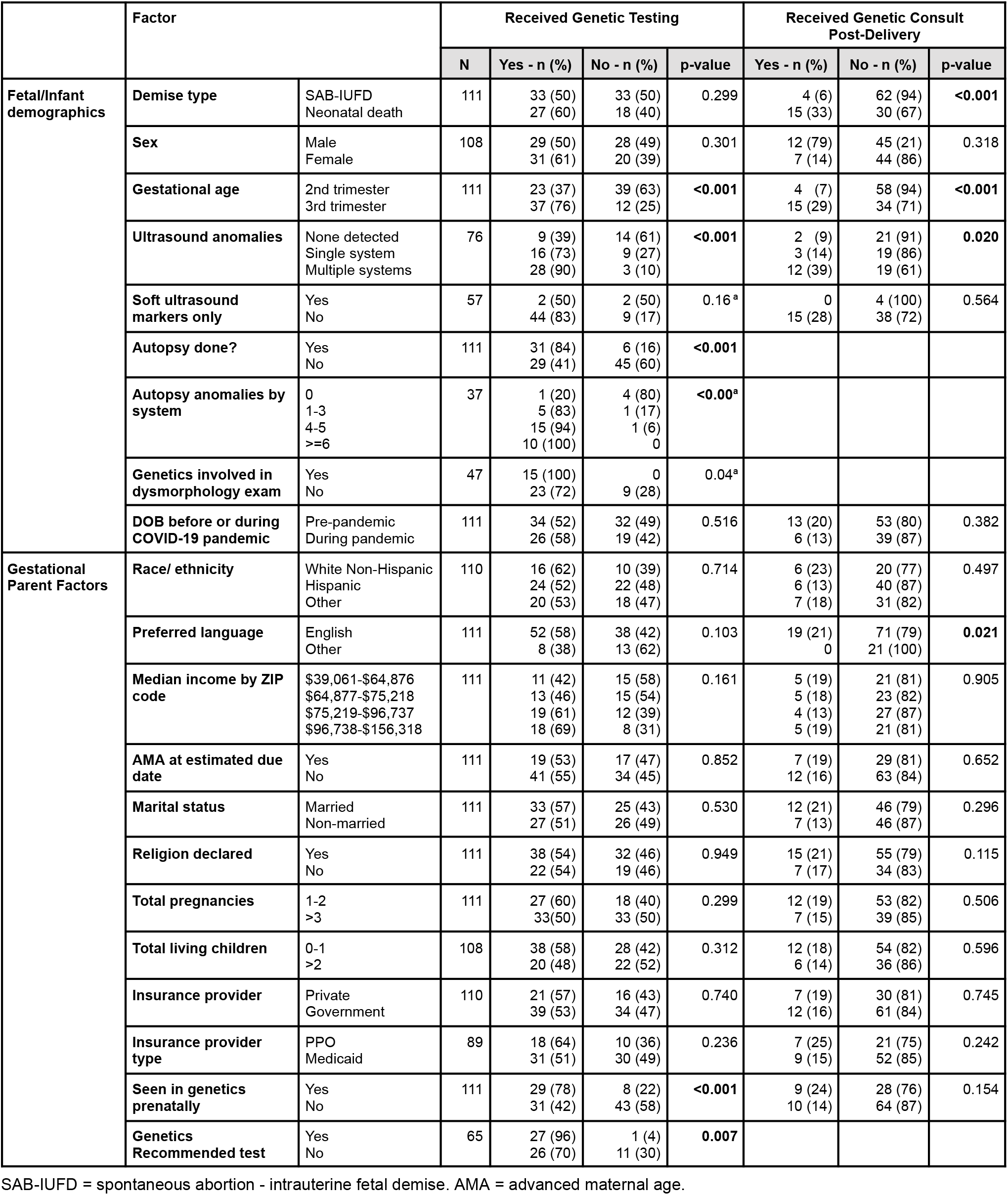
Comparison of fetal/infant characteristics and reception of genetic testing and/or genetics consult.

|  | Factor |  | Received Genetic Testing |  |  |  | Received Genetic Consult Post-Delivery |  |  |
| --- | --- | --- | --- | --- | --- | --- | --- | --- | --- |
|  |  |  | N | Yes - n (%) | No - n (%) | p-value | Yes - n (%) | No - n (%) | p-value |
| **Fetal/Infant demographics** | **Demise type** | SAB-IUFD<br>Neonatal death | 111 | 33 (50)<br>27 (60) | 33 (50)<br>18 (40) | 0.299 | 4 (6)<br>15 (33) | 62 (94)<br>30 (67) | **<0.001** |
|  | **Sex** | Male<br>Female | 108 | 29 (50)<br>31 (61) | 28 (49)<br>20 (39) | 0.301 | 12 (79)<br>7 (14) | 45 (21)<br>44 (86) | 0.318 |
|  | **Gestational age** | 2nd trimester<br>3rd trimester | 111 | 23 (37)<br>37 (76) | 39 (63)<br>12 (25) | **<0.001** | 4 (7)<br>15 (29) | 58 (94)<br>34 (71) | **<0.001** |
|  | **Ultrasound anomalies** | None detected<br>Single system<br>Multiple systems | 76 | 9 (39)<br>16 (73)<br>28 (90) | 14 (61)<br>9 (27)<br>3 (10) | **<0.001** | 2 (9)<br>3 (14)<br>12 (39) | 21 (91)<br>19 (86)<br>19 (61) | **0.020** |
|  | **Soft ultrasound markers only** | Yes<br>No | 57 | 2 (50)<br>44 (83) | 2 (50)<br>9 (17) | 0.16<sup>a</sup> | 0<br>15 (28) | 4 (100)<br>38 (72) | 0.564 |
|  | **Autopsy done?** | Yes<br>No | 111 | 31 (84)<br>29 (41) | 6 (16)<br>45 (60) | **<0.001** |  |  |  |
|  | **Autopsy anomalies by system** | 0<br>1-3<br>4-5<br>>=6 | 37 | 1 (20)<br>5 (83)<br>15 (94)<br>10 (100) | 4 (80)<br>1 (17)<br>1 (6)<br>0 | **<0.00<sup>a</sup>** |  |  |  |
|  | **Genetics involved in dysmorphology exam** | Yes<br>No | 47 | 15 (100)<br>23 (72) | 0<br>9 (28) | 0.04<sup>a</sup> |  |  |  |
|  | **DOB before or during COVID-19 pandemic** | Pre-pandemic<br>During pandemic | 111 | 34 (52)<br>26 (58) | 32 (49)<br>19 (42) | 0.516 | 13 (20)<br>6 (13) | 53 (80)<br>39 (87) | 0.382 |
| **Gestational Parent Factors** | **Race/ ethnicity** | White Non-Hispanic<br>Hispanic<br>Other | 110 | 16 (62)<br>24 (52)<br>20 (53) | 10 (39)<br>22 (48)<br>18 (47) | 0.714 | 6 (23)<br>6 (13)<br>7 (18) | 20 (77)<br>40 (87)<br>31 (82) | 0.497 |
|  | **Preferred language** | English<br>Other | 111 | 52 (58)<br>8 (38) | 38 (42)<br>13 (62) | 0.103 | 19 (21)<br>0 | 71 (79)<br>21 (100) | **0.021** |
| | **Median income by ZIP code** | \$39,061-\$64,876<br>\$64,877-\$75,218<br>\$75,219-\$96,737<br>\$96,738-\$156,318 | 111 | 11 (42)<br>13 (46)<br>19 (61)<br>18 (69) | 15 (58)<br>15 (54)<br>12 (39)<br>8 (31) | 0.161 | 5 (19)<br>5 (18)<br>4 (13)<br>5 (19) | 21 (81)<br>23 (82)<br>27 (87)<br>21 (81) | 0.905 |
|  | **AMA at estimated due date** | Yes<br>No | 111 | 19 (53)<br>41 (55) | 17 (47)<br>34 (45) | 0.852 | 7 (19)<br>12 (16) | 29 (81)<br>63 (84) | 0.652 |
|  | **Marital status** | Married<br>Non-married | 111 | 33 (57)<br>27 (51) | 25 (43)<br>26 (49) | 0.530 | 12 (21)<br>7 (13) | 46 (79)<br>46 (87) | 0.296 |
|  | **Religion declared** | Yes<br>No | 111 | 38 (54)<br>22 (54) | 32 (46)<br>19 (46) | 0.949 | 15 (21)<br>7 (17) | 55 (79)<br>34 (83) | 0.115 |
|  | **Total pregnancies** | 1-2<br>>3 | 111 | 27 (60)<br>33(50) | 18 (40)<br>33 (50) | 0.299 | 12 (19)<br>7 (15) | 53 (82)<br>39 (85) | 0.506 |
|  | **Total living children** | 0-1<br>>2 | 108 | 38 (58)<br>20 (48) | 28 (42)<br>22 (52) | 0.312 | 12 (18)<br>6 (14) | 54 (82)<br>36 (86) | 0.596 |
|  | **Insurance provider** | Private<br>Government | 110 | 21 (57)<br>39 (53) | 16 (43)<br>34 (47) | 0.740 | 7 (19)<br>12 (16) | 30 (81)<br>61 (84) | 0.745 |
|  | **Insurance provider type** | PPO<br>Medicaid | 89 | 18 (64)<br>31 (51) | 10 (36)<br>30 (49) | 0.236 | 7 (25)<br>9 (15) | 21 (75)<br>52 (85) | 0.242 |
|  | **Seen in genetics prenatally** | Yes<br>No | 111 | 29 (78)<br>31 (42) | 8 (22)<br>43 (58) | **<0.001** | 9 (24)<br>10 (14) | 28 (76)<br>64 (87) | 0.154 |
|  | **Genetics Recommended test** | Yes<br>No | 65 | 27 (96)<br>26 (70) | 1 (4)<br>11 (30) | **0.007** |  |  |  |

SAB-IUFD = spontaneous abortion - intrauterine fetal demise. AMA = advanced maternal age.

### 3.2 Characteristics of anatomy US

Eighty perinates had anatomy US available for review. Of these 80 cases, 66% had anomalies in one or more systems, 5% had soft markers only (e.g. two-vessel cord), while the remaining 29% had a normal US (Figure 3).

**Figure 3.**
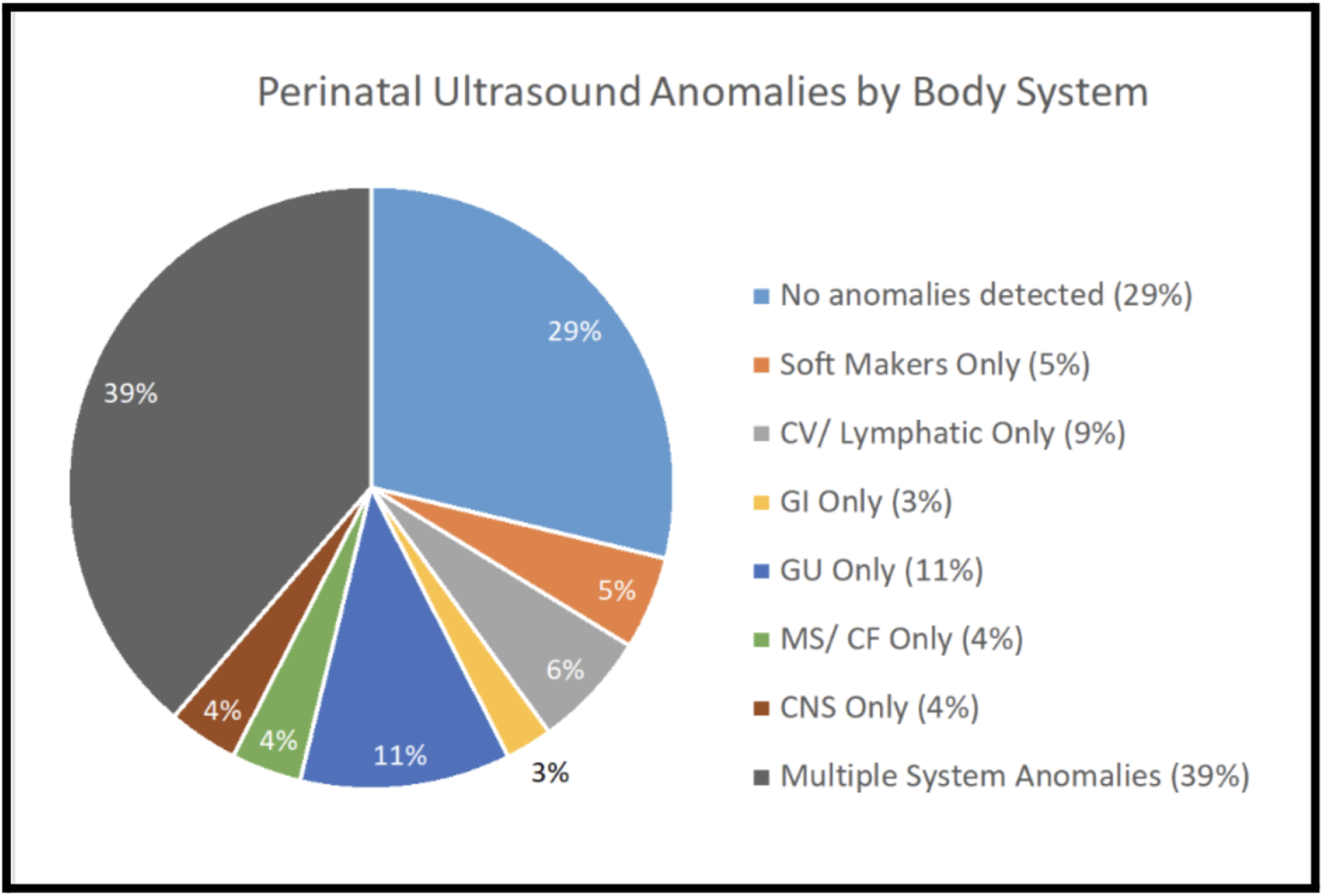
Anatomy US anomalies by body system (N=80). CNS = Central nervous system, CV = Cardiovascular; GI Gastrointestinal; GU = Genitourinary; MS/CF = Musculoskeletal/ Craniofacial

### 3.3 Geneticist involvement

Forty-six (41.4%) perinatal cases had documented referrals to prenatal genetics and 37 (33.3%) attended. Twenty-eight (25.2%) perinates were referred for genetics consult post-delivery and 19 (17.1%) received one. The cases that were referred but did not receive an inpatient consult were seen outpatient or had already received negative results. Overall only 46 patients were seen prenatally, postnatally, or both (41%), despite the fact that 66% (73 cases) of all demise cases had US abnormalities. Forty-seven (42.3%) cases received a dysmorphology exam. The exam was performed by a medical geneticist (in 31.9% of the 47 who had any type of dysmorphology exam), pathologist, obstetrician, or neonatologist.

### 3.4 Genetic testing

Sixty (54.0%) perinates (out of 111) received at least one type (most commonly chromosomal microarray analysis, CMA) of genetic testing prenatally or postnatally. Seven additional cases were recommended to have genetic testing, but the family declined. Three patients were recommended to have WES or WGS, and one received WGS. Of the 60 perinates that received genetic testing, 24 (40.0%) received a diagnostic result (pathogenic or likely pathogenic), 4 (6.7%) received a variant of uncertain significance (VUS), 1 (1.7%) received a result of sex discrepancy, and 31 (51.7%) received a negative result. Of the 24 perinates that received a diagnostic result, 11 (45.8%) had a monogenic disorder, 10 (41.7%) had a chromosomal aneuploidy, 2 (8.3%) had a disorder caused by a copy number variant, and 1 (4.1%) had genome-wide uniparental isodisomy (Supplemental Information Table 2).

**Table 2.** Comparison of fetal/infant and gestational parent demographic characteristics and diagnostic yield.

|  | Factor |  | Received Abnormal Result |  |  |  |
| --- | --- | --- | --- | --- | --- | --- |
|  |  |  | N | Yes - n (%) | No - n (%) | p-value |
| **Fetal/Infant Factors** | **Demise type** | SAB-IUFD<br>Neonatal death | 60 | 13 (39)<br>16 (56) | 20 (61)<br>11 (44) | 0.114 |
|  | **Sex** | Male<br>Female | 60 | 13 (45)<br>16 (52) | 16 (55)<br>15 (48) | 0.599 |
|  | **Gestational age** | 2nd trimester<br>3rd trimester | 60 | 9 (39)<br>20 (54) | 14 (61)<br>17 (46) | 0.261 |
|  | **Ultrasound anomalies** | None detected<br>Single system<br>Multiple systems | 53 | 1 (11)<br>7 (44)<br>18 (64) | 8 (89)<br>9 (56)<br>10 (36) | **0.016** |
|  | **Soft ultrasound markers only** | Yes<br>No | 46 | 1 (50)<br>25 (57) | 1 (50)<br>19 (43) | 1.000 |
|  | **Autopsy anomalies by system** | 0<br>1-3<br>4-5<br>≥6 | 31 | 0<br>2 (40)<br>9 (60)<br>3 (30) | 1(100)<br>3 (60)<br>6 (40)<br>7 (70) | 0.371 |
|  | **Genetics involved in dysmorphology exam** | Yes<br>No | 38 | 10 (67)<br>8 (35) | 5 (33)<br>15 (65) | 0.054 |
|  | **Date of birth before or during COVID-19 pandemic** | Pre-pandemic<br>During pandemic | 60 | 16 (47)<br>13 (50) | 18 (53)<br>13 (50) | 0.798 |
| **Gestational Parent Factors** | **Race/ ethnicity** | White Non-Hispanic<br>Hispanic<br>Other | 60 | 7 (44)<br>12 (50)<br>9 (45) | 9 (56)<br>12 (50)<br>11 (55) | 0.759 |
|  | **Preferred language** | English<br>Other | 60 | 26 (50)<br>3 (38) | 26 (50)<br>5 (63) | 0.510 |
| | **Median income by ZIP code** | \$39,061-\$64,876<br>\$64,877-\$75,218<br>\$75,219-\$96,737<br>\$96,738-\$156,318 | 60 | 5 (45)<br>9 (69)<br>6 (33)<br>8 (50) | 6 (55)<br>4 (31)<br>12 (67)<br>10 (50) | 0.267 |
|  | **Advanced maternal age at estimated due date** | Yes<br>No | 60 | 13 (68)<br>15 (36) | 6 (32)<br>27 (64) | **0.034** |
|  | **Marital status** | Married<br>Non-married | 60 | 18 (55)<br>11 (41) | 15 (46)<br>16 (59) | 0.287 |
|  | **Religion declared** | Yes<br>No | 60 | 19 (50)<br>10 (46) | 19 (50)<br>12 (55) | 0.734 |
|  | **Total pregnancies** | 1-2<br>>3 | 60 | 11 (41)<br>18 (47) | 16 (59)<br>15 (53) | 0.622 |
|  | **Total living children** | 0-1<br>>2 | 58 | 16 (42)<br>12 (60) | 22 (58)<br>8 (40) | 0.195 |
|  | **Insurance provider** | Private<br>Government | 60 | 11 (52)<br>18 (46) | 10 (48)<br>21 (54) | 0.645 |
|  | **Seen in genetics prenatally** | Yes<br>No | 60 | 17 (59)<br>12 (38) | 12 (41)<br>20 (63) | 0.123 |
|  | **Genetics recommended testing** | Yes<br>No | 53 | 20 (74)<br>6 (23) | 7 (26)<br>20 (77) | **<0.001** |

SAB-IUFD = spontaneous abortion - intrauterine fetal demise.

Of the 60 cases that underwent genetic testing, 35 (58.3%) were documented as discussed with the gestational parent. Documentation of discussion was more common if results were abnormal (22 of 29 abnormal results, 75.9%). The result reporting rate was not statistically different between genetics and non-genetics providers.

### 3.5 Autopsy

Autopsy discussion was documented in 97 cases, 37 of which consented and underwent autopsy. The remainder were undecided, declined by a parent, or declined by the coroner’s office (one case). Of the 37 autopsies performed, 13.5% (5 cases) had no anomalies on autopsy and the remainder had one or more. This is summarized in Supplemental Material Table 7. Cases of neonatal death received more autopsies than SAB-IUFD cases (p=0.001). No other significant relationships between autopsy uptake and case characteristics were detected.

### 3.6 Predicting referral to genetics and uptake of genetic testing

Cases involving English speaking families (p<0.021) and neonatal demise (p<0.001) received more genetics consults than non-English speaking families and SAB-IUFD cases, respectively. However, neither demise type nor language spoken were shown to significantly affect the rate of genetic testing. More genetics consults (p<0.001) and testing (p<0.001) were done in cases with third trimester deliveries (compared to second trimester). A greater number of US abnormalities was also associated with increased frequency of genetic consults (p=0.020) and genetic testing (p<0.001). Cases in which prenatal genetics consults were done (p<0.001), autopsies were done (p<0.001), autopsies identified abnormalities (p<0.001), and/or genetics offered the testing (as compared to other specialists, p=0.007) were all significantly more likely to have had genetic testing done. These factors did not appear to increase rates of post-delivery genetics consult. No statistically significant relationship was found between the remaining factors. Results are summarized in Table 1.

### 3.7 Predicting abnormal genetic testing results

Perinate and gestational parent factors were compared with results from prenatal and postnatal genetic testing. More anomalies on prenatal US (p=0.016), geneticist involvement in the dysmorphology exam (p=0.028), team recommending testing (genetics vs. other, p<0.001), and advanced maternal age (AMA, p=0.046) were all associated with abnormal genetic testing results. Results are summarized in Table 2.

### 3.8 Results return and counseling

Of the 60 perinatal demises that received genetic testing, result return was documented in the UCI EMR in 35 cases. Abnormal results were more likely to be returned (or be documented as returned) than normal results (p=0.029) amongst all provider types. Seven abnormal results did not have documentation of result return in the UCI EMR. These included 3 cases in which testing was recommended by non-genetics providers (all with a VUS) and 3 cases in which testing was recommended by genetics providers, all of which were lost to follow up. More patient education points were documented as discussed by genetics than by non-genetics providers (p<0.001) (Table 3).

**Table 3.**
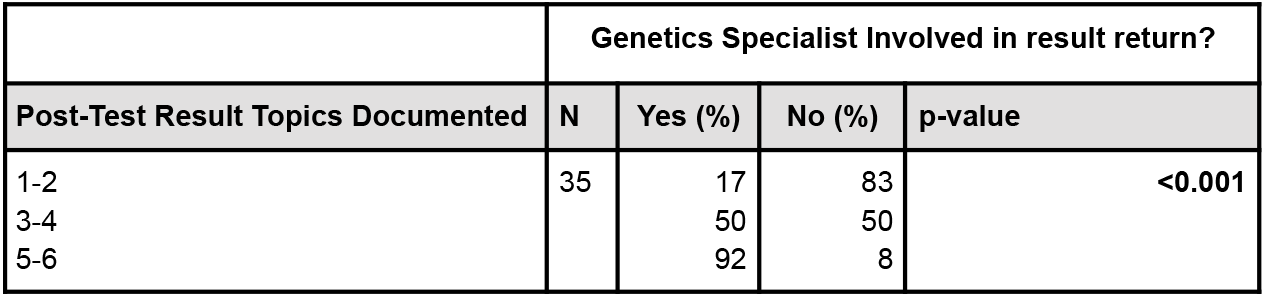
Number of post-test patient education topics documented.

## Discussion

The plateauing decline of the perinatal death rate is an ongoing challenge in public health. Undiagnosed genetic conditions are thought to contribute to a significant number deaths among perinates both with and without major congenital anomalies.^14,15^ ACOG currently recommends genetic evaluation for all cases of perinatal demise, but genetic evaluation is not always straightforward and isn’t always offered when it should be.^16^ The main goal of this study was to investigate factors that affect the rate of genetic consultations, rate of genetic testing, and diagnostic yield of that testing in hopes that the findings could spur the development of a more clearly defined process for handling, understanding, and ultimately preventing perinatal demise. The type and trimester of demise, number of US anomalies, occurrence of and number of anomalies detected on autopsy, occurrence of a geneticist-performed dysmorphology exam, language spoken, and prenatal and/or postnatal genetics specialist involvement were all associated with significant changes in the rate of genetic consults, rate of genetic testing, and/or diagnostic yield of genetic testing included in this study.

Not surprisingly, the number of prenatal US anomalies was positively associated with a higher rate of genetic consultations (p = 0.020), rate of genetic testing (p < 0.001), and diagnostic yield of genetic testing (p = 0.016). Similarly, deceased perinates who received an autopsy were more likely to receive genetic testing (p < 0.001), and a greater number of autopsy abnormalities was associated with more frequent genetic testing (p < 0.001). It is important to note that there was no increase in consultations, testing, or yield in cases with only soft markers, though there were only four such cases in this cohort. Interestingly, a higher rate of autopsy was noted in neonatal demise than in SAB-IUFDs (p = 0.001), but there was no significant difference between gestational age at delivery for autopsy uptake (p=0.137). This finding suggests that whether the perinate is born living may impact autopsy uptake more than gestational age at delivery, though provider differences in obtaining autopsy consent can’t be excluded.

Families with English listed as their preferred language received significantly more consultations (p = 0.021) than families with less English proficiency - in fact, none of the 21 non-English speaking families received postnatal genetics consultations. The rate of genetic disease is not lower in medically underserved populations like those who speak a different language, but unfortunately this trend in consultation rates is not novel despite availability of interpretation services.^17^ These findings merit further study regarding potential barriers to consultation for both non-English speaking patients and for consultants. We analyzed many other factors which were not significantly associated with the rate of consultations, testing, or yield in this study. On initial analysis, advanced maternal age (age 35 years or older at time of delivery) seemed to be associated with a higher diagnostic yield (p = 0.034). However, when chromosomal abnormalities (known increase with advanced maternal age) were removed from the analysis, there was no significant association, consistent with the literature.^18^ Of note, in our cohort median income, insurance type, and insurance provider did not independently affect referral rates.

Neonatal demise cases received significantly more genetic consultations than SAB-IUFD cases (p<0.001), despite pre-existing data showing that the rate of genetic anomalies is similar in SAB-IUFDs (13.7%) and neonatal demise cases (13.6% in hospitalized infants).^5, 19^ Why this occurred in this study is unknown. Perhaps the time available between birth and death in neonatal demise cases simply provides greater opportunity for consultation. Neonatologist involvement and identification of abnormalities not seen prenatally may provide further impetus for consultation. However, if neonatologist involvement and postnatal exam were the cause for greater consult rate in neonatal demise, we might also expect higher rates of genetic testing in neonatal demise (vs. SAB-IUFD), but this was *not* observed. It is possible this study lacked the power to determine a difference, neonates were too ill for genetic testing, or geneticists were more selective with testing recommendations when a more accurate postnatal dysmorphology exam was possible. If time and additional workup increase the consult rate, it follows that third trimester demise cases would also have an increased number of genetic consultations (p<0.001). Third trimester cases were also more likely to receive genetic testing (p<0.001), despite the fact that congenital anomalies cause a higher percentage of stillbirths in the second trimester than the third.^19^ This could again relate to timing - in the third trimester families are more likely to have received a consultation, may have had more time to contemplate and complete testing, and may be more motivated to pursue testing in a fetus who is still living.

Though the tumult of the COVID-19 pandemic led to significant reductions in antenatal and postnatal care attendance country-wide (appointment changes/cancellations estimated at 53.4%), no changes in the rate of genetic testing, consultation, or diagnostic yield were observed in our cohort.^20,21,22^ In fact, preliminary data from other studies specifically demonstrated efficient conversion of genetic consultations into telehealth format during the pandemic.^23^ In theory, a virtual consult format may enable a single geneticist to perform more consults when a detailed, in person dysmorphology exam is not required. Combining this information with our data, which indicate that time may be an important factor influencing consult rate, perhaps converting some consultations into a virtual format could increase consultation rate and therefore result in a greater number of genetic diagnoses for deceased perinates.

Overall diagnostic yield in our cohort was about 47%. The rate of genetic testing and diagnostic yield was higher when a genetic specialist recommended the test (p = 0.007 and p <0.001, respectively). Similarly, genetic testing occurred more in cases when a geneticist performed a dysmorphology exam (p = 0.042) and the difference in diagnostic yield approached significance (p = 0.054). The importance of the dysmorphology exam, which geneticists specialize in, can not be overstated - it guides differential diagnosis, test selection, and result interpretation. In one study, a geneticist’s dysmorphology exam and pathologist’s histological workup were together able to identify a definitive genetic diagnosis in 31% and a suggested diagnosis in 22% of cases prior to confirmatory testing.^24^ Genetics involvement can also help ensure that appropriate testing is ordered.

One study found that 26% of genetic testing orders placed by other professionals were corrected to include more appropriate testing after order review by genetic counselors.^25^ Genetics professional involvement likely also improves family and patient education. More patient education points (p < 0.001) were documented by Geneticists for both normal and abnormal result returns (though we can’t rule out a difference in documentation completeness). Thirdly, genetics professionals can improve continuity of care by clearly documenting and explaining genetic testing results.^26^ For example, this study identified three cases with abnormal genetic testing results which would not have caused the perinate’s physical anomalies or demise. These results could have implications for family members, but should not mark the end of perinatal demise investigation. These are complicated situations in which in-depth genetic counseling, follow up, and possibly further genetic testing would be needed.

There are some important limitations of this study to note. Firstly, record access was at times challenging when outside facilities were involved. This may have limited the power of our data collection and influenced results. Secondly, it is likely that cases for which genetics was consulted were more likely to be affected by genetic disease, thus exacerbating any observed increase in diagnostic yield among the group that received consultations. Finally, this study embarked on a primarily univariate analysis. However, some factors such as attendance to genetics consult and occurrence of a dysmorphology exam carry the possibility of an overlapping effect on diagnostic outcomes. An extension of this study could use multivariate analysis to identify predictors of outcomes of genetics evaluation. Further studies may also benefit from a larger sample size to better determine differences between groups. Future research should include consideration of testing type, standardization of ICD-10 coding to facilitate study, efficiency of consultation workflow, effects of genetic consult timing related to the demise, and the thoroughness of post-test counseling and results return.

## Conclusions

In summary, the type of perinatal demise (neonatal > SAB-IUFDs), the timing of demise (third > second trimester), number of anomalies on US and/or autopsy, and primary language were all significantly associated with changes in the rates of genetic consultations, genetic testing, and/or diagnostic yield in this cohort. Though the influence of ascertainment bias could not be eliminated, this cohort taught us that the combined effort of multiple specialists, including the genetics team, likely improves the care of these perinates and their families. Proponents of the multidisciplinary approach have cited a lack of access to genetics professionals as a barrier to these collaborative arrangements.^27^ Indeed, in our study more genetics consults were performed when more time was available to complete them. Increasing the availability of genetics professionals by use of virtual consults or other time saving measures may thus improve multidisciplinary patient care. Regardless of improvements in consultant availability and/or efficiency, not all cases require Genetics involvement, and therefore all providers should aim to improve the rate of genetic testing to include all perinatal demise cases. Perhaps a national or even institutional guideline could encourage this as well as facilitate research to identify new candidate causative genes.^28^ Even perinatal demise in the case of abnormal fetomaternal circulation (e.g. thrombus, abruption) may have genetic underpinnings and should be offered testing.^29^ One group studying WES yield followed their cohort longitudinally and found that families who received diagnostic genomic autopsy utilized this knowledge for subsequent pregnancy planning. Five of these families underwent IVF with pre-implantation genetic testing and subsequently had healthy babies.^14^ Understanding genetic contributions to perinatal demise is essential for outcome improvement, and the first step is ensuring that all perinatal demise cases are offered genetic testing.

## Data Availability

All data produced in the present study are available upon reasonable request to the authors

## Acknowledgements

We would like to thank Dr. Robert Edwards, MD and Pamela Aron Johnson, RN for their help with identification of the patient cohort.

## References

1. Barfield WD, Committee on Fetus and Newborn, Watterberg K et al. Standard terminology for fetal, infant, and perinatal deaths. Pediatrics May 2016;137(5):e20160551.

2. CDC Stillbirth. Data and Statistics on Stillbirth 2025. https://www.cdc.gov/stillbirth/data-research/index.html#:∼:text=Stillbirth%20affects%20about%201%20in,the%20first%20year%20of%20life. Accessed January 21, 2026.

3. Dongarwar D, Aggarwal A, Barning K, Salihu HM. Trends in stillbirths and stillbirth phenotypes in the United States: an analysis of 131.5 million births. International Journal of Maternal and Child Health and AIDS 2020;9:146.

4. Unicef. Levels and Trends in Child Mortality 2023. https://data.unicef.org/resources/levels-and-trends-in-child-mortality. Accessed January 21, 2026.

5. Stillbirth Collaborative Research Network Writing Group. Causes of death among stillbirths. JAMA 2011;306(22):2459–2468.

6. Wapner RJ. Genetics of stillbirth. Clinical Obstetrics & Gynecology 2010;53(3):628–634.

7. Wilders R. Cardiac ion channelopathies and the sudden infant death syndrome. ISRN Cardiology 2012;846171.

8. American College of Obstetricians and Gynecologists. Management of stillbirth: Obstetric care consensus no. 10. Obstetrics and Gynecology 2020;135(3):e110–e132.

9. Giordano JL, Wapner RJ. Genomics of stillbirth. Semin Perinatol 2024;48(1):151866.

10. Burton JL, Underwood J. Clinical, educational, and epidemiological value of autopsy. The Lancet 2007;369(9571):1471–1480.

11. Hutchinson JC, Potocki L, Van den Veyver IB. Current controversies in prenatal diagnosis 2: Conventional postmortem examination remains the gold standard for the anatomical examination of fetal loss. Prenatal Diagnosis 2025;45(10):1343–1350.

12. United States Census Bureau. American Community Survey Data 2025. https://www.census.gov/programs-surveys/acs/data.html. Accessed January 21, 2026.

13. Baker DL, Eash T, Schuette JL, Uhlmann WR. Guidelines for Writing Letters to Patients. J Genet Couns. 2002;11(5):399–418.

14. Byrne AB, Arts P, Ha TT, Kassahn KS, Pais LS, O’Donnell-Luria A. Genomic autopsy to identify underlying causes of pregnancy loss and perinatal death. Nat Med. 2023;29(1):180–189.

15. Yang X, Bai R, Zhang J et al. Analysis of the Causes of Neonatal Death and genetic variations in congenital anomalies: a multi-center study. Front. Pediatr. 2024;12:1419495.

16. Marouane A, Olde Keizer RACM, Frederix GWJ, Vissers LELM, de Boode WP, van Zelst-Stams WAG. Congenital anomalies and genetic disorders in neonates and infants: A single-center observational cohort study. European Journal of Pediatrics 2022;181(1):359–367.

17. Muessig KR, Zepp JM, Keast E et al. Retrospective assessment of barriers and access to genetic services for hereditary cancer syndromes in an integrated health care delivery system. Hereditary Cancer in Clinical Practice 2022;20(1):7.

18. Gardner RJM, Amor DJ. Gardner and Sutherland’s chromosome abnormalities and genetic counseling (Fifth edition). Oxford University Press 2018.

19. Liu L, Huang H, Yu M, Su H. Analysis of intrauterine fetal demise: A hospital-based study in Taiwan over a decade. Taiwanese Journal of Obstetrics and Gynecology 2013;52(4):546–550.

20. Preis H, Mahaffey B, Heiselman C, Lobel M. Vulnerability and resilience to pandemic-related stress among U.S. women pregnant at the start of the COVID-19 pandemic. Social Science & Medicine 2020;266:113348.

21. Khalil A, Von Dadelszen P, Draycott T, Ugwumadu A, O’Brien P, Magee L. Change in the incidence of stillbirth and preterm delivery during the COVID-19 pandemic. JAMA 2020;324(7), 705–706.

22. Shah PS, Ye XY, Yang J, Campitelli MA. Preterm birth and stillbirth rates during the COVID-19 pandemic: A population-based cohort study. CMAJ 2021;193(30):E1164–E1172.

23. Mann C, Goodhue B, Guillard A et al. The COVID-19 pandemic and reproductive genetic counseling: Changes in access and service delivery at an academic medical center in the United States. Journal of Genetic Counseling 2021;30(4), 958–968.

24. Aggarwal S, Tandon A, Das Bhowmik A, Safarulla JMNJ, Dalal A. A dysmorphology based systematic approach toward perinatal genetic diagnosis in a fetal autopsy series. Fetal and Pediatric Pathology 2018;37(1), 49–68.

25. Miller CE, Krautscheid P, Baldwin EE et al. Genetic counselor review of genetic test orders in a reference laboratory reduces unnecessary testing. American Journal of Medical Genetics Part A 2014;164(5), 1094–1101.

26. Hunt Brendish K, Patel D, Yu K et al. Genetic counseling clinical documentation: Practice resource of the national society of genetic counselors. Journal of Genetic Counseling 2021;30(5), 1336–1353.

27. Pasquier L, Minguet G, Moisdon-Chataigner S et al. How do non-geneticist physicians deal with genetic tests? A qualitative analysis. European Journal of Human Genetics 2022;30(3), 320–331.

28. Dolanc Merc M, Peterlin B, Lovrecic L. The genetic approach to stillbirth: A systematic review. Prenatal Diagnosis 2023;43(9):1220–1228.

29. Rosner M, Kolbe T, Hengstschläger M. Fetomaternal microchimerism and genetic diagnosis: On the origins of fetal cells and cell-free fetal DNA in the pregnant woman. Mutation Research 2021;788:108399

