## Supplemental Information for "Investigating Uptake and Impact of Genetic and Genomic Evaluation Following Perinatal Demise"

### Table 1. Demographics of gestational parents

| Demographic | | n = 111 | % |
|---|---|---|---|
| **Race/ ethnicity** | Asian<br>Black or African American<br>Hispanic White<br>Non-Hispanic White<br>One or more<br>Unknown/ Missing/ Prefers not to declare | 12<br>7<br>46<br>26<br>19<br>1 | 10.8<br>6.3<br>41.4<br>23.4<br>17.1<br>0.9 |
| **Preferred Language** | English<br>Spanish<br>Vietnamese<br>Arabic<br>Portuguese | 90<br>17<br>2<br>1<br>1 | 81.1<br>15.3<br>1.8<br>0.9<br>0.9 |
| **Marital Status** | Married<br>Non-Married | 58<br>53 | 52.3<br>47.7 |
| **Religious Affiliation** | Baptist<br>Buddhist<br>Catholic<br>Christian<br>The Church of Jesus Christ of Latter-day Saints<br>Evangelical<br>Jehovah’s Witness<br>Muslim<br>Non-Denominational Christian<br>Non-Religious<br>Other<br>Unknown/ Missing/ Prefers not to declare | 2<br>1<br>41<br>19<br>3<br>1<br>2<br>1<br>1<br>15<br>1<br>24 | 1.8<br>0.9<br>36.9<br>17.1<br>2.7<br>0.9<br>1.8<br>0.9<br>0.9<br>13.5<br>0.9<br>21.6 |
| **Insurance Type** | Medicaid<br>PPO<br>HMO<br>Tricare<br>EPO<br>Not Documented | 71<br>28<br>8<br>2<br>1<br>1 | 64.0<br>25.2<br>7.2<br>1.8<br>0.9<br>0.9 |
| **Age at delivery, years** | Mean (S.D.)<br>Median<br>Range | 32 (5)<br>32<br>18-42 | |
| **Total number of pregnancies** | Mean (S.D.)<br>Median<br>Range | 3 (2)<br>3<br>1-14 | |
| **Total Number of Living Children** | Mean (S.D.)<br>Median<br>Range | 1 (1)<br>1<br>0-6 | |
| **Median family income by ZIP code (dollars)** | Mean (S.D.)<br>Median<br>Range | $81,290 (24,139)<br>$75,219<br>$39,061 – 156,318 | |

### Table 2. Cases that received genetic diagnosis

| Genetic Diagnostic Category for Pathogenic/ Likely Pathogenic Results | Number of Perinatal Demise Cases, N = 24 | % | Genetic Diagnostic Testing Type |
|---|---|---|---|
| **Chromosomal Conditions** | **n = 10** | **41.7** | |
| Trisomy 13 | 1 | | Karyotype |
| Mosaic Trisomy 13 | 1 | | SNP CMA |
| Trisomy 18 | 3 | | Karyotype, FISH |
| Trisomy 21 | 3 | | Karyotype |
| Triple X Syndrome | 1 | | Karyotype |
| Triploidy | 1 | | CMA |
| **Copy Number Variant Disorders** | **n = 2** | **8.3** | |
| 16p11.2 Duplication Syndrome | 1 | | CMA |
| 16p11.2 Deletion Syndrome | 1 | | CMA |
| **Uniparental Disomy Disorders** | **n = 1** | **4.1** | |
| Beckwith-Wiedemann Syndrome | 1 | | SNP CMA[a] |
| **Monogenic Conditions** | **n = 11** | **45.8** | |
| Autosomal Recessive Polycystic Kidney Disease | 2 | | Panel |
| COL2A1-Related Disorder | 1 | | Panel |
| Hemoglobin Barts Syndrome | 1 | | NBS[b], Panel |
| Nager Syndrome | 1 | | CMA |
| Noonan-Spectrum Syndrome | 2 | | Panel, ES |
| Osteogenesis Imperfecta | 1 | | Panel |
| Syndromic Microphthalmia | 1 | | Panel |
| Thanatophoric Dysplasia | 2 | | Panel |

**a** = CMA revealed genome-wide uniparental isodisomy mosaicism. Beckwith-Wiedemann Syndrome and Russell Silver Syndrome Methylation by MLPA was normal. Assumption of BWS diagnosis based on clinical findings; **b** = One case of Hemoglobin Barts Syndrome was diagnosed initially by the California Newborn Screening Program (NBS) and confirmed by an alpha thalassemia panel. CMA = Chromosomal microarray; ES = Exome sequencing; FISH = Fluorescence in situ hybridization.

### Table 3. Demographics of perinates

| Demographics (Categorical) | | n = 111 | % |
|---|---|---|---|
| **Demise Type** | SAB-IUFD<br>Neonatal Deaths | 66<br>45 | 59.5<br>40.5 |
| **Sex** | Male<br>Female<br>Not Documented | 57<br>51<br>3 | 51.4<br>45.9<br>2.7 |
| **Was Demise a Part of a Multiple Gestation?** | Yes<br>No | 11<br>100 | 9.9<br>90.1 |
| **Demographics (Continuous)** | | | |
| **Gestational Age (weeks)** | Mean (S.D.)<br>Median<br>Range | 27 (6)<br>26<br>20-41 | |
| **Birth Weight (grams)** | Mean (S.D.)<br>Median<br>Range | 1,310.08 (1,043.70)<br>832.50<br>190-4,320 | |

**Table 4. Anatomy ultrasound (U/S) demographics**

| Characteristic | | N | n | % |
|---|---|---|---|---|
| **Received First Anatomy U/S** | Yes<br>No/ Not Documented | 111 | 80<br>31 | 72.1<br>27.9 |
| **Gestation Age at First Anatomy U/S** | 1st Trimester<br>2nd Trimester<br>3rd Trimester | 80 | 42<br>36<br>2 | 52.5<br>85.7<br>4.8 |
| **Number of Ultrasound System Anomalies** | None<br>Soft markers only<br>Anomalies in a single system<br>Anomalies in multiple systems | 80 | 23<br>4<br>22<br>31 | 28.8<br>5.0<br>31.2<br>38.8 |
| **Infants with Anomalies in a Single System** | Respiratory System<br>Cardiovascular/ Lymphatic System<br>Gastrointestinal System<br>Reticuloendothelial System<br>Genitourinary System<br>Musculoskeletal/ Craniofacial System<br>Endocrine System<br>Central Nervous System | 22 | 0<br>5<br>2<br>0<br>9<br>3<br>0<br>3 | 0<br>22.7<br>9.1<br>0<br>40.9<br>13.6<br>0.0<br>13.6 |
| **Presence of Soft Markers** | No anomalies detected<br>Soft markers only<br>Soft markers and non-soft markers<br>Only non-soft markers | 80 | 23<br>4<br>20<br>33 | 28.8<br>5.0<br>25.0<br>41.3 |
| **Ultrasound Clinic Location** | UCIMC<br>External | 80 | 40<br>40 | 50.0<br>50.0 |
| **Ultrasound Reading Physician** | MFM<br>Non-MFM | 80 | 66<br>14 | 82.5<br>17.5 |

**Table 5. Demographics of cases referred for prenatal and postnatal genetics evaluations.**

| Characteristic | | N | n | % |
|---|---|---|---|---|
| **Referred to genetics prenatally?** | Yes<br>No | 111 | 46<br>65 | 41.4<br>58.5 |
| **Attended prenatal genetics visit?** | Yes<br>No | 111 | 37<br>74 | 33.3<br>66.7 |
| **Prenatal genetics clinic affiliation** | UCIMC<br>Non-UCI | 37 | 26<br>11 | 70.3<br>29.7 |
| **Recommended genetics consult after delivery by a genetic counselor?** | Yes<br>No | 37 | 12<br>25 | 32.4<br>67.6 |
| **Referred to genetics consult postdelivery?** | Yes<br>No | 111 | 28<br>83 | 25.2<br>74.8 |
| **Received postdelivery genetics consult?** | Yes<br>No | 111 | 19<br>92 | 17.1<br>82.9 |
| **Medical geneticists involved in dysmorphology exam?** | Yes<br>No<br>NA, did not receive dysmorphology exam | 111 | 15<br>32<br>64 | 13.5<br>28.8<br>57.7 |
| **Received at least one genetic test prenatally and/or postnatally?** | Yes<br>No | 111 | 60<br>51 | 54.1<br>45.9 |

**Table 6. Summary of perinatal cases that did not attend post-delivery consult with medical geneticist after being referred for consult following delivery (N=9)**

| Referral Indication for Genetics Consult | Genetic Testing (Recommended by) | Genetic Testing Received | Follow-Up | Referred to Genetics Post-Result? | Seen in Genetics at UCI Post-Referral? |
|---|---|---|---|---|---|
| IUGR | Normal CMA (OB) | At Delivery | OB RR | No | N/A |
| IUFD | Mosaic Trisomy 13 by CMA (OB) | At Delivery | OB RR, referred to GC | Yes | No |
| IUFD | 16p11.2 duplication syndrome by CMA (OB) | At Delivery | RR by OB, referral to GC | Yes | No |
| Pregnancy loss at 20 weeks | Triploidy by CMA (Genetics, OB)[a] | At Delivery | RR by OB, referred to GC | Yes | No |
| Severe hydrops | CMA: VUS 11p15.4 deletion<br>Panel: SPTA1 c.5572C>G<br>Panel: Pathogenic HBA1/HBA2 variant (Genetics)[b] | At delivery | Genetics phone consult only; RR by NICU, declined GC | Yes | No, declined |
| Tetralogy of Fallot, hydrops | Normal CMA, postnatal (Pediatric cardiology) | At Delivery | Not documented | No | N/A |
| Suspected skeletal dysplasia | FGFR3 c.746C>G [pathogenic], FGFR3 c.2417C>T [VUS] (Genetics)[c] | At delivery | RR by GC | Yes | Yes |
| Megacystis | Normal CMA (Neonatology) | Prenatally | RR by neonatology | No | N/A |
| Fetal cardiac anomaly | Normal CMA (Neonatology) | At delivery | PPV not attended | No | N/A |

a = Offered prenatally by a genetic counselor, but gestational parent elected to test products of conception after delivery. Obstetrics recommended CMA testing again after delivery. b = recommended by phone consult with medical geneticist; c = offered prenatally by a genetic counselor, but gestational parent elected to test neonate after delivery. CMA = Chromosomal microarray; GC = Genetic counselor; IUFD = Intrauterine fetal demise; IUGR = Intrauterine growth restriction; PPV = post-partum visit; SNP = Single nucleotide polymorphism; VUS = Variant of uncertain significance. RR = Result Return, CMA = chromosomal microarray

**Table 7. Comparison of fetal/infant and gestational parent demographic characteristics between perinatal demise subjects who received autopsy and those who did not.**

| Factor | | Received Autopsy | | | |
|---|---|---|---|---|---|
| **Fetal/ Infant Demographics** | | **N** | **Yes - n (%)** | **No - n (%)** | **p-value** |
| **Demise type** | SAB-IUFD<br>Neonatal death | 111 | 14 (21.1%)<br>23 (51.1%) | 52 (78.8%)<br>22 (48.9%) | 0.001 |
| **Sex** | Male<br>Female | 108 | 19 (33.3%)<br>18 (35.3%) | 38 (66.7%)<br>33 (64.7%) | 0.830 |
| **Gestational age at delivery** | Second trimester<br>Third trimester | 111 | 17 (27.4%)<br>20 (40.8%) | 45 (72.6%)<br>29 (59.2%) | 0.137 |
| **Ultrasound anomalies** | None detected<br>Single system<br>Multiple systems | 76 | 8 (34,8%)<br>7 (31.8%)<br>16 (51.6%) | 15 (65.2%)<br>15 (68.2%)<br>15 (48.4%) | 0.275 |
| **DOB before or during COVID-19 pandemic** | Pre-pandemic<br>During pandemic | 111 | 23 (34.8%)<br>14 (31.1%) | 43 (65.2%)<br>31 (68.9%) | 0.682 |
| **Gestational Parent Demographics** | | | | | |
| **Race/ ethnicity** | White Non-Hispanic<br>Hispanic<br>Other | 110 | 10 (38.5%)<br>16 (34.8%)<br>11 (28.9%) | 16 (61.5%)<br>30 (65.2%)<br>27 (71.1%) | 0.714 |
| **Preferred language** | English<br>Other | 111 | 32 (35.5%)<br>5 (23.8%) | 58 (64.4%)<br>16 (76.2%) | 0.304 |
| **Median income by ZIP code** | $39,061-$64,876<br>$64,877-$75,218<br>$75,219-$96,737<br>$96,738-$156,318 | 111 | 8 (32.0%)<br>6 (21.4%)<br>12 (38.7%)<br>10 (38.5%) | 17 (68.0%)<br>22 (78.6%)<br>19 (61.3%)<br>16 (61.5%) | 0.472 |
| **AMA at EDD** | Yes<br>No | 111 | 11 (30.6%)<br>26 (34.7%) | 25 (69.4%)<br>49 (65.3%) | 0.667 |
| **Marital status** | Married<br>Non-married | 111 | 17 (29.3%)<br>20 (37.7%) | 41 (70.7%)<br>33 (62.3%) | 0.347 |
| **Religion declared** | Yes<br>No | 111 | 23 (32.9%)<br>14 (34.1%) | 47 (67.1%)<br>27 (65.8%) | 0.889 |
| **Total Pregnancies** | 1-2<br>>3 | 111 | 17 (37.8%)<br>20 (30.3%) | 28 (62.2%)<br>46 (69.7%) | 0.412 |
| **Total living children** | 0-1<br>>2 | 108 | 26 (39.4%)<br>11 (26.2%) | 40 (60.6%)<br>31 (73.8%) | 0.159 |
| **Insurance provider category** | Private<br>Government | 110 | 12 (32.4%)<br>25 (34.2%) | 25 (67.6%)<br>48 (65.8%) | 0.849 |
| **Insurance provider type** | PPO<br>Medicaid | 89 | 11 (39.3%)<br>19 (31.1%) | 17 (60.7%)<br>42 (68.9%) | 0.451 |
| **Seen in genetics prenatally** | Yes<br>No | 111 | 13 (35.1%)<br>24 (32.4%) | 24 (64.9%)<br>50 (67.6%) | 0.776 |

**Table 8. Summary of perinatal cases seen prenatally who did not receive recommended postnatal consult (N=6).**

| Referral Indication for Prenatal GC | Genetic Testing (Recommended by) | Genetic Testing Received | Follow-Up |
|---|---|---|---|
| U/S finding of multiple fetal anomalies | Normal karyotype; Medicaid insurance denied reflex to CMA (Obstetrics) | Prenatally | Result returned by GC |
| U/S finding of hydrops | None, declined | None | N/A |
| AMA, U/S findings of anhydramnios, scoliosis, and clubfoot | Sex discrepancy identified by CMA; (Genetic counselor) | Following delivery | Result returned by GC; gestational parent sample to r/o MCC |
| U/S findings of micromelia, talipes, and pulmonary hypoplasia | Thanatophoric dysplasia by panel; (Genetic counselor) | Following delivery | Result returned by GC |
| Suspected fetal skeletal dysplasia | COL2A1-related disorder by panel (Genetic counselor) | Following delivery | Result returned by GC |
| U/S finding of fetal abnormality, AMA | Trisomy 21 by CMA (External) | Prenatally | Result returned by GC |

GC = Genetic counselor; MCC = maternal cell contamination; R/o = rule out; U/S = ultrasound
